# Frequency of antimicrobial-resistant bloodstream infections in Thailand, 2022

**DOI:** 10.1101/2024.06.01.24308013

**Authors:** Krittiya Tuamsuwan, Panida Chamawan, Phairam Boonyarit, Voranadda Srisuphan, Preeyarach Klaytong, Chalida Rangsiwutisak, Prapass Wannapinij, Trithep Fongthong, John Stelling, Paul Turner, Direk Limmathurotsakul

**Affiliations:** The Office of Permanent Secretary, Ministry of Public Health, Nonthaburi, 11000, Thailand; Mahidol Oxford Tropical Medicine Research Unit, Faculty of Tropical Medicine, Mahidol University, Bangkok, Thailand; Brigham and Women’s Hospital and Harvard Medical School, Boston, MA, United States; Cambodia-Oxford Medical Research Unit, Angkor Hospital for Children, Siem Reap, Cambodia; Centre for Tropical Medicine and Global Health, Nuffield Department of Medicine, University of Oxford, Oxford, United Kingdom; Department of Tropical Hygiene, Faculty of Tropical Medicine, Mahidol University, Bangkok, Thailand

**Keywords:** Antimicrobial resistant, surveillance, bloodstream infection, proportion, incidence, frequency

## Abstract

**Objectives:** To evaluate the frequency of antimicrobial-resistant bloodstream infections (AMR BSI) in Thailand

**Methods:** We analyzed data from 2022, generated by 111 public hospitals in health regions 1 to 12, using the AutoMated tool for Antimicrobial resistance Surveillance System (AMASS) and submitted to the Ministry of Public Health, Thailand. Multilevel Poisson regression models were used.

**Results:** The most common cause of community-origin AMR BSI was third-generation cephalosporin-resistant *Escherichia coli* (3GCREC, 65.6%; 5,101/7,773 patients) and of hospital-origin AMR BSI was carbapenem-resistant *Acinetobacter baumannii* (CRAB, 51.2%, 4,968/9,747 patients). The percentage of patients tested for BSI was negatively associated with the frequency of community-origin 3GCREC BSI and hospital-origin CRAB BSI. Hospitals in health regions 4 (lower central) had the highest frequency of community-origin 3GCREC BSI per 100,000 tested patients (adjusted incidence rate ratio, 2.06; 95% confidence interval: 1.52-2.97). Health regions were not associated with the frequency of hospital-origin CRAB BSI, although between-hospital variation was high even adjusting for hospital level and size.

**Conclusion:** The high between-hospital variation of hospital-origin CRAB BSI suggests the importance of hospital-specific factors. Our approach and findings highlight health regions and hospitals where actions against AMR infection, including antimicrobial stewardship and infection control, should be prioritized.

**Highlights:**

- The frequency of AMR BSI in 111 public hospitals in Thailand in 2022 was studied.
- The frequency of community-origin 3GCREC BSI was different by regions.
- The frequency of hospital-origin CRAB BSI varied greatly among hospitals.
- Underuse of BC was associated with the higher frequency of AMR BSI per tested patients.
- Our findings contributed to actions against AMR at local and national levels.

## Introduction

Strengthening the knowledge and evidence base through surveillance and research is one of the five key objectives outlined in the global action plan on antimicrobial resistance (AMR), led by the World Health Organization (WHO) [1]. At the global level, the WHO Global Antimicrobial Resistance and Use Surveillance System (GLASS) has provided a standard approach to the collection, analysis and sharing of data [2]. The GLASS country profile dashboard presents summaries of AMR data (including AMR proportions and AMR frequency) that each country reported to the system [3]. In addition, many high-income countries (HICs), such as England, have collated and made local AMR data available via a publicly accessible web tool [4]. Better access, use and research on surveillance data have been one of the main elements to tackle AMR [4–6].

In 2020, we developed the AutoMated tool for Antimicrobial resistance Surveillance System (AMASS), an offline application that allow hospitals to automatically analyse and generate standardized antimicrobial resistance (AMR) surveillance reports from their routine microbiology and hospital data [7]. This tool was designed to overcome the limitations of inadequate human resources to analyze AMR data and generate summary reports in low and middle-income countries (LMICs). The automatically generated reports stratify infection origin into community-origin and hospital-origin based on the recommendations of WHO GLASS [2] and provide additional reports on mortality (%) and total number of deaths following AMR and non-AMR bloodstream infections (BSI). We initially tested the application in seven hospitals in seven countries [7]. We recently reported the difference of frequency of hospital-origin AMR BSI between tertiary-care hospitals (TCHs) and secondary-care hospitals (SCHs) using the AMASS and a retrospective data in 49 public hospitals from 2012 to 2015 [8]. From December 2022 to January 2023, on behalf of the Health Administration Division, Ministry of Public Health (MoPH) Thailand, we trained 25 public hospitals in Thailand to utilize the AMASS, and reported the benefit of local and timely AMR data review [9].

Following the success of the early implementation of the AMASS in 25 public hospitals [9], in 2023 the MoPH invited all 127 public referral hospitals in Thailand to utilize the AMASS using their data from 2022 and submit the summary data to the MoPH. This study aims to evaluate the frequency of community-origin and hospital-origin AMR BSI in Thailand.

## Materials and Methods

### Study setting

In 2022, Thailand had a population of 66.1 million, consisted of 77 provinces, and covered 513,120 km^2^. The health systems in each province were integrated into 12 groups of provinces, known as health regions, plus the capital Bangkok as health region 13 (Figure 1A), using the concept of decentralization [10]. The Health Administration Division, Ministry of Public Health (MoPH) supervised 127 public referral hospitals in health regions 1 to 12. These included 35 advanced-level referral hospital (i.e. level A, with a bed size of about 500-1,200), 55 standard-level referral hospital (i.e. level S, with a bed size of about 300-500) and 37 mid-level referral hospital (i.e. level M1, with a bed size of about 180-300) [11]. All level A and S hospitals, and most of level M1 hospitals were equipped with a microbiology laboratory capable of performing bacterial culture using standard methodologies for bacterial identification and susceptibility testing provided by the Department of Medical Sciences, MoPH.

**Figure 1.**
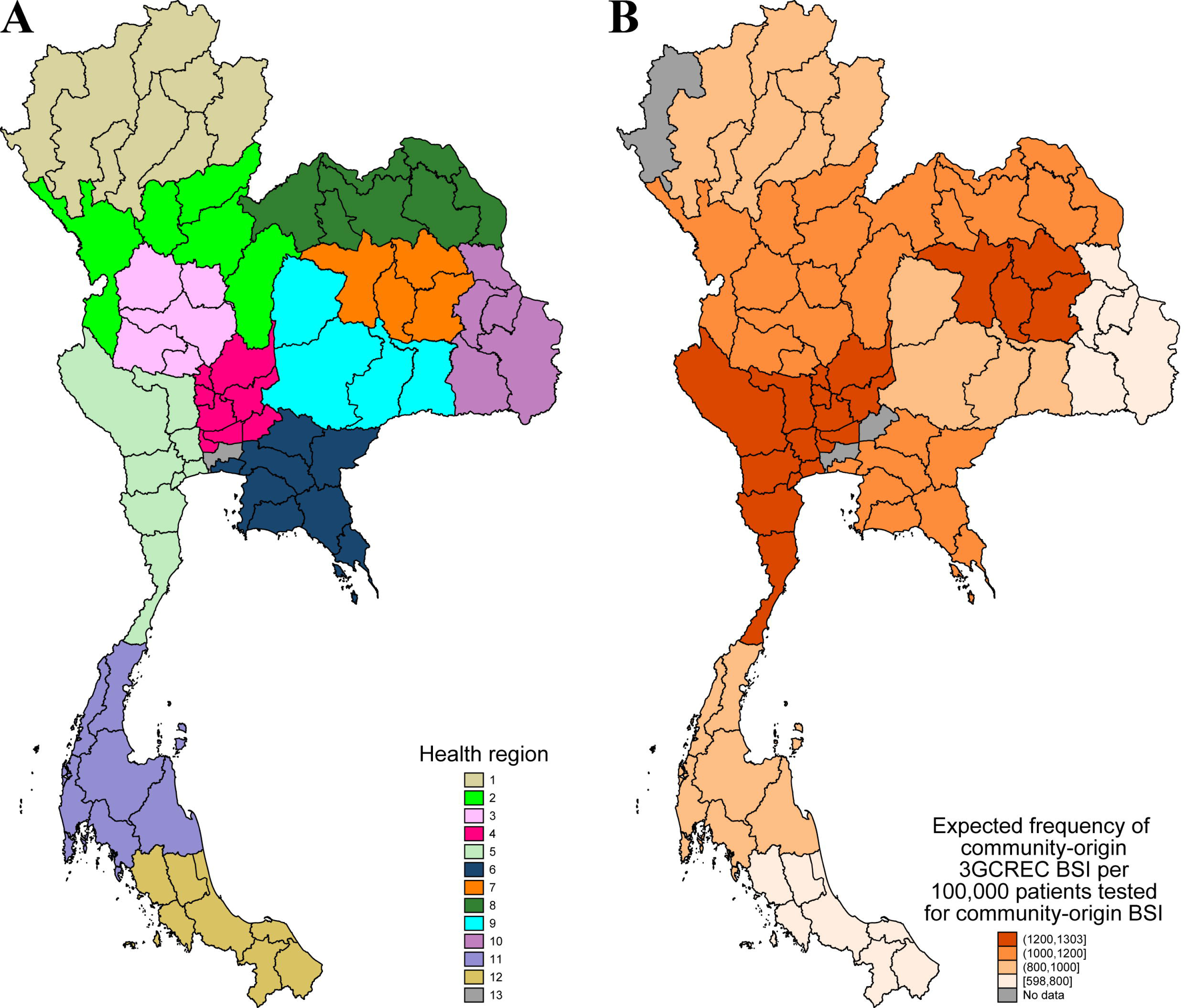
Health regions in Thailand (1A) and expected frequency of community-origin 3GCREC BSI (1B) by health regions. Footnote of figure 1. For each health region, the expected frequency was estimated based on the total number of expected cases per the total number of patients tested for BSI in the provinces. The number of expected cases was calculated using the fixed effects of the covariates in the final model and assuming that all hospitals had the percentage of patients having BC taken within the first 2 days of hospital admission at the mean value (16.9%).

### Study tool and its implementation

From 16 December 2022 to 30 June 2023, on behalf of the Health Administration Division, MoPH, we invited and trained staff from 127 public referral hospitals in health regions 1 to 12 to utilize the AMASS with their own microbiology and hospital admission data files via four online meetings, five face-to-face meetings and on-line support. Then, the MoPH invited all 127 public referral hospitals to utilize the AMASS in 2023 using their data from 2022. The additional two online meetings for 127 hospitals were conducted in January 2023, and the four face-to-face meetings were conducted in Chonburi (for hospitals in health regions 4, 5 and 6), Chiang Mai (for hospitals in health regions 1, 2 and 3), Surajthani (for hospitals in health regions 11 and 12) and Khon Kaen (for hospital in health regions 7, 8, 9 and 10) provinces in January and February 2023.

The content of the online meetings and face-to-face meetings were as previously described [9]. In short, we taught the hospitals how to use the AMASS, complete the data dictionary files, and verify and read the AMR reports automatically generated by the AMASS. The hospitals were request to validate the reports using two methods: (a) checking data verification log files generated the AMASS whether all information was imported accurately (e.g. total number of specimens, total number of hospital admissions, number of missing values, total number of isolates per organism in the raw microbiology data file, total number of antibiotics being tested, and all pathogens and antimicrobial susceptibility testing results under evaluation), and (b) comparing the summary data and reported generated by the AMASS with data generated from manual calculations obtained from complete line listing of several organisms. The hospitals were asked to re-export the data files (if needed), complete the data dictionary files, re-run the AMASS and verify the reports until the reports were accurate. The 25 hospitals who were initially trained in the first round [9] supported the other hospitals in their health regions.

Then, the hospitals who completed the utilization of the AMASS submitted the summary data generated by the AMASS to the MoPH. A new interactive Thailand AMR dashboard was launched in 2023 and is open-access (Figure 2) [12].

**Figure 2.**
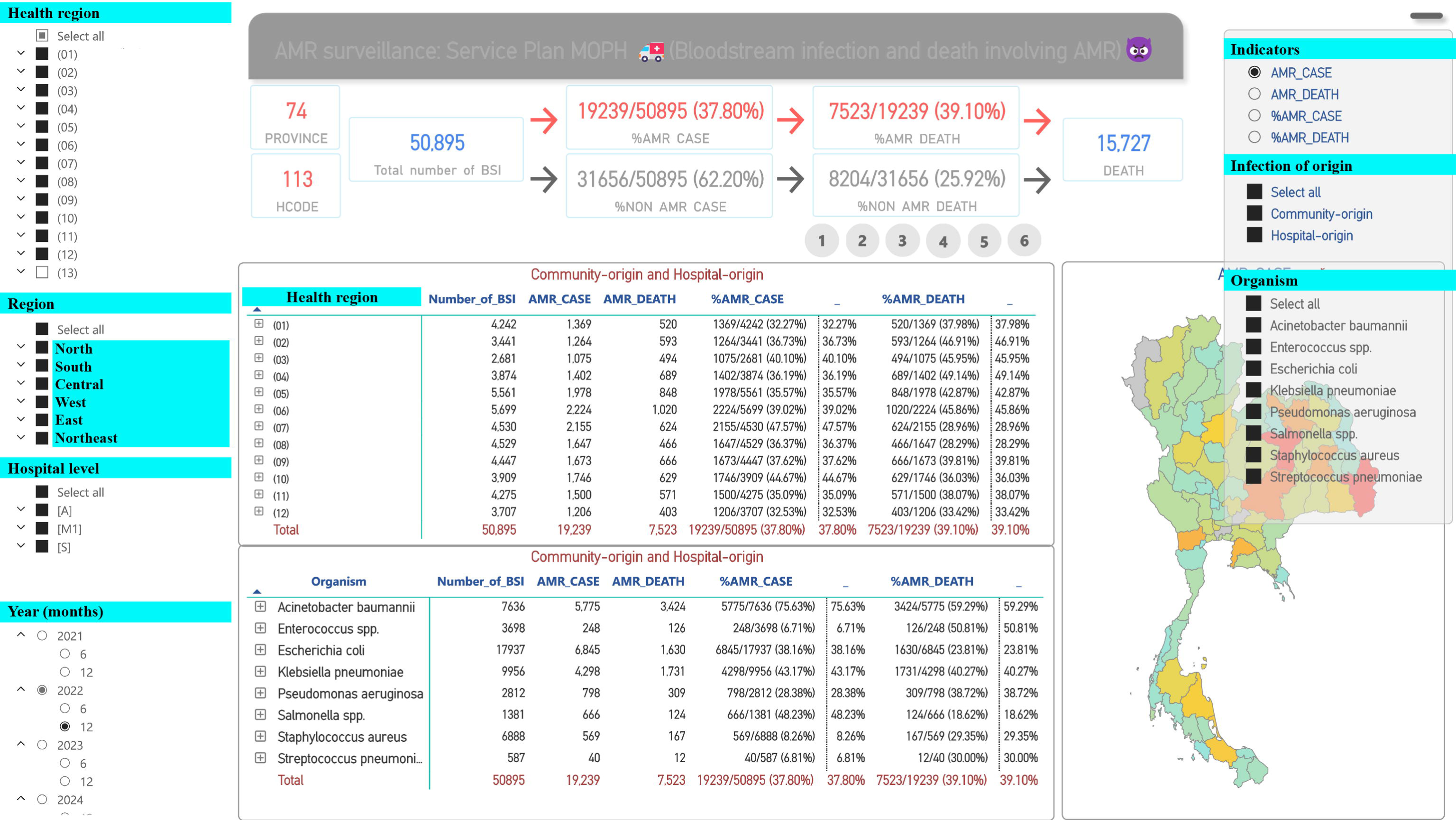
New interactive Thailand AMR dashboard [12]. Footnote of figure 2. Thai words were translated to English and highlighted in green color. The picture was captured from the website [12] on 30 June 2023.

### Study design

This is a retrospective study evaluating frequency of AMR BSI across public referral hospitals in Thailand using the hospital-level AMR data submitted to the MoPH. For AMR infections, we initially analyzed the following organisms: carbapenem-resistant *Acinetobacter baumannii* (CRAB), carbapenem-resistant *Pseudomonas aeruginosa* (CRPA), carbapenem-resistant *Escherichia coli* (CREC), carbapenem-resistant *Klebsiella pneumoniae* (CRKP), third-generation cephalosporin-resistant *E. coli* (3GCREC) and *K. pneumoniae* (3GCRKP), and methicillin-resistant *Staphylococcus aureus* (MRSA) which are in the WHO GLASS priority list of AMR bacteria and are of locally importance. Only blood culture (BC) results were included in the analysis of the AMASS and in this study. The AMASS de-duplicated the data by selecting only the first admission of a patient with BSI caused by a given bacterial species per patient per reporting period and per infection origin.

### Definitions

Community-origin BSI was defined for patients with first positive blood specimens in the hospital taken within the first two calendar days of admission with calendar day one equal to the day of admission [13]. Patients with first positive blood specimens taken after the first two calendar days were categorized as cases of hospital-origin BSI. The classification of community-origin and hospital-origin BSI was performed within the AMASS and based on specimen collection dates and hospital admission dates extracted from the microbiology and hospital data files of each hospital. Blood specimens collected from patients who were not admitted at the study hospitals were excluded from the analysis. These specimens were categorized as BSI of unknown origin in the summary reported generated by the AMASS [7].

Patients who had the first BC taken within the first two calendar days of hospital admission were considered as having tested for community-origin BSI. Patients were considered at risk for hospital-origin BSI after they stayed in the hospital for more than two calendar days. Patients who had the first BC taken after the first two calendar days of hospital admission were considered as having tested for hospital-origin BSI.

The proportion of AMR (%) was calculated as the percentage of patients with new AMR BSI over all patients with new BSIs for each pathogen during the reporting period. The frequency of AMR BSI for each pathogen was calculated as the total number of new patients with AMR BSI during the reporting period per 100,000 tested patients, per 100,000 admissions and per 100,000 patient-days at risk for hospital-origin BSI.

Mortality (%) following AMR BSI for each pathogen under evaluation was calculated as the percentage of patients with new AMR BSI who died in the hospital, using all-cause in-hospital mortality in the discharge summary regularly completed by the attending physicians and available in the admission data of the hospitals.

### Statistical analysis

Medians, interquartile ranges (IQR) and ranges of continuous variables were estimated. IQRs are presented in terms of 25th and 75th percentiles. For percentages, one digit after the decimal points is shown when the total number of denominators is higher than 100. To evaluate BC utilization, we calculated percentage of patients tested for BSI and rate of patients tested for BSI per 1,000 bed-days. We did not calculate the total number of BC specimens per 1,000 bed-days [14, 15], because, during the workshops, we observed that some hospitals recorded two BC specimens as two separate records, while some hospitals recorded those as a single record in their laboratory information systems (LIS). Therefore, we utilized the deduplicated number of patients tested for BSI as the indicator of BC utilization. Kruskal Wallis test was used to compare the median of continuous variables between groups.

We evaluated factors associated with the frequency of community-origin 3GCREC BSI and hospital-origin CRAB BSI using univariable and multivariable Poisson random-effects regression models. The community-origin 3GCREC BSI and hospital-origin CRAB BSI were selected as they were the most common causes of community-origin and hospital-origin AMR BSI, respectively. The random effect model accounts for the fact that the observed outcomes for some hospitals can take on extreme values because of random variation. Factors evaluated included health region, hospital level, hospital bed size, Gross Provincial Product (GPP), rate of patients having BC taken per 1,000 patient-days, percentage of patients having BC taken within the first two calendar days of hospital admission (for community-origin AMR BSI) and percentage of patients having BC taken after first two calendar days of hospital admission (for hospital-origin AMR BSI). Data of GPP of Thailand in 2021 was used as a proxy for the size of the economy in each province [16]. We summarized results with incidence rate ratio (IRRs) and 95% confidence intervals (Cis).

To measure heterogeneity in the multilevel Poisson regression, we calculated the median rate ratio (MRR) and variance partition coefficient (VPC) using the methods described by Austin et al [17] (Appendix A). The MRR is the median relative change in the rate of the occurrence of the event when comparing two hospitals (i.e. two clusters) with identical covariates from two randomly selected different hospitals. The VPC denotes the proportion of the (unexplained) variation in the outcome that is beyond that explained by the fixed effects of the covariates in the models. In other words, the VPC denotes the proportion of the variation that is due to between-hospital (i.e. between-cluster) variation [17]. We calculated the VPC using exact calculations [17] and total number of tested patients for 111 covariate patterns of the 111 hospitals included in the study. We used STATA (version 14.2; College Station, Texas) for all analyses.

### Data availability

The hospital-level summary data used for the study are open-access and available at https://figshare.com/s/6aee792c15290d7dea3c.

### Ethics

Ethical permission for this study was obtained from the Institute for the Committee of the Faculty of Tropical Medicine, Mahidol University (TMEC 23-085). Individual patient consent was not sought as this was a retrospective study using summary data, and the Ethical and Scientific Review Committees approved the process.

## Results

### Baseline characteristics

Of 127 public referral hospitals, 116 (91%) used the AMASS to analyze their microbiology and hospital admission data files, and submitted the summary AMR surveillance reports and data from 2022 to the MoPH. Four hospitals had no data of BC with ‘no growth’ results in the electronic data file, and one hospital exported and used inaccurate hospital admission data. These hospitals were removed from the analysis because the data of ‘no growth’ specimens and accurate data of hospital admissions were required for calculating the frequency of AMR BSI. Therefore, a total of 111 hospitals were included in the final analysis.

Of all public referral hospitals in Thailand, 100% of Level A hospitals (35/35), followed by 89% of Level S hospitals (49/55) and 73% of Level M1 hospitals (27/37) were included in this study. The data were available from 74 of 77 provinces (96%) in Thailand, all provinces except Mae Hong Son, Nakorn Nayok and Bangkok.

From 1 January to 31 December 2022, there were 3,916,766 hospital admissions of 3,042,979 patients, totaling 23,675,466 patient-days, across 111 public referral hospitals included in the study. Hospital characteristics are presented in Table 1 and S1. A total of 618,187 patients had BC taken, resulting in 20.3% of patients having BC taken (618,187/3,024,360). Rate of patients having BC taken was 26.1 per 1,000 patient-days (618,187/23,541,908).

**Table 1.**
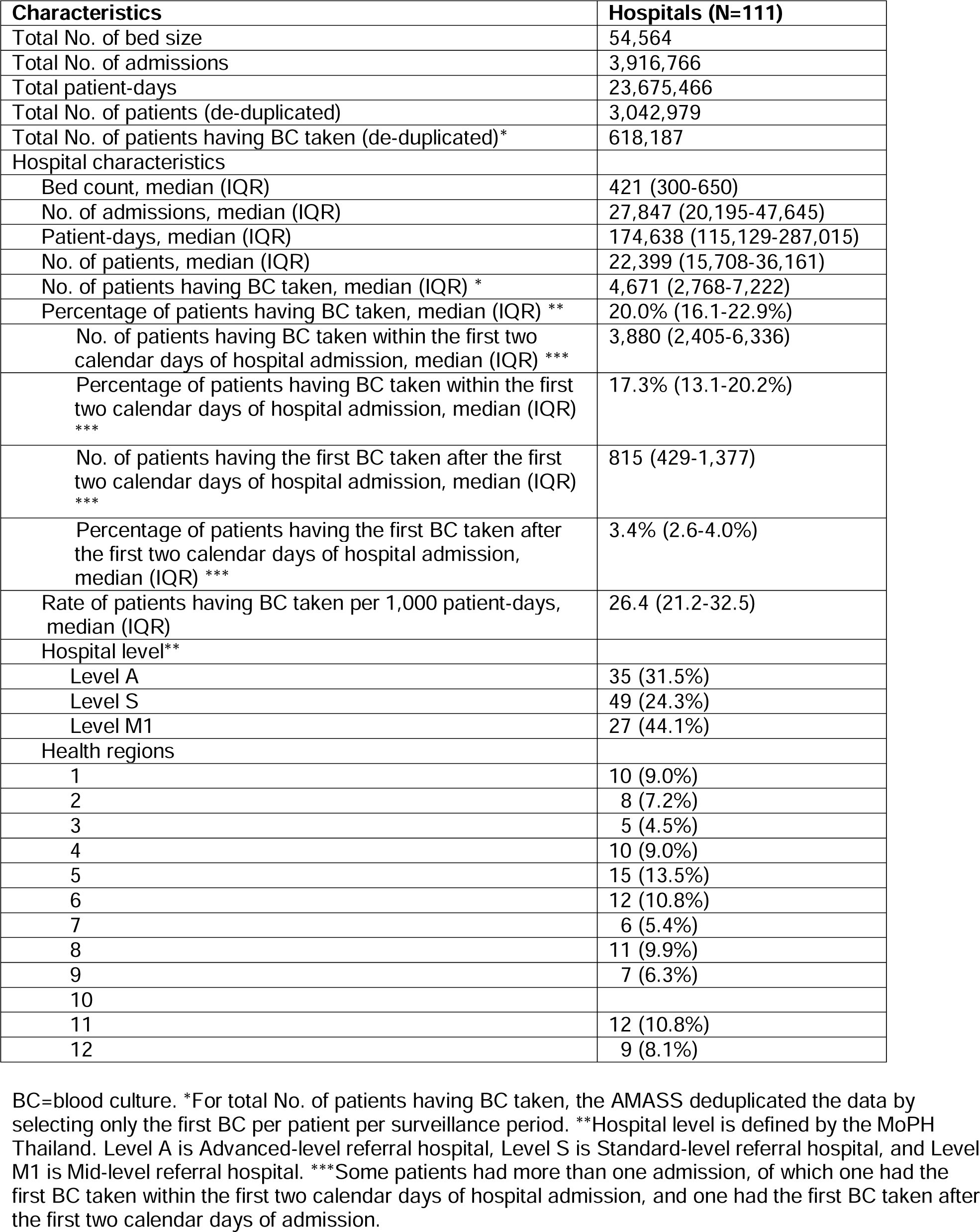
Characteristics of 111 public referral hospitals in Thailand, 2022.

The median percentage of patients having BC taken within the first two calendar days of hospital admission was highest in Level A hospitals (18.8%), followed by that in Level S (16.5%) and Level M1 (15.0%) hospitals (p=0.11). The median percentage of patients having the first BC taken after the first two calendar days of hospital admission was also highest in Level A hospitals (4.2%), followed by that in Level S (3.3%) and Level M1 (2.6%) hospitals (p<0.001).

### Total No. of cases and deaths following AMR BSI

There were 7,829 patients with community-origin AMR BSI caused by the pathogens under evaluation (Table 2). The most common cause of community-origin AMR BSI was 3GCREC, accounting 65.9% (5,157/7,829) of those patients. The total number of deaths following community-origin 3GCREC BSI was 1,048, representing 54.1% (1,048/1,936) of all deaths following community-origin AMR BSI caused by pathogens under evaluation.

**Table 2.**
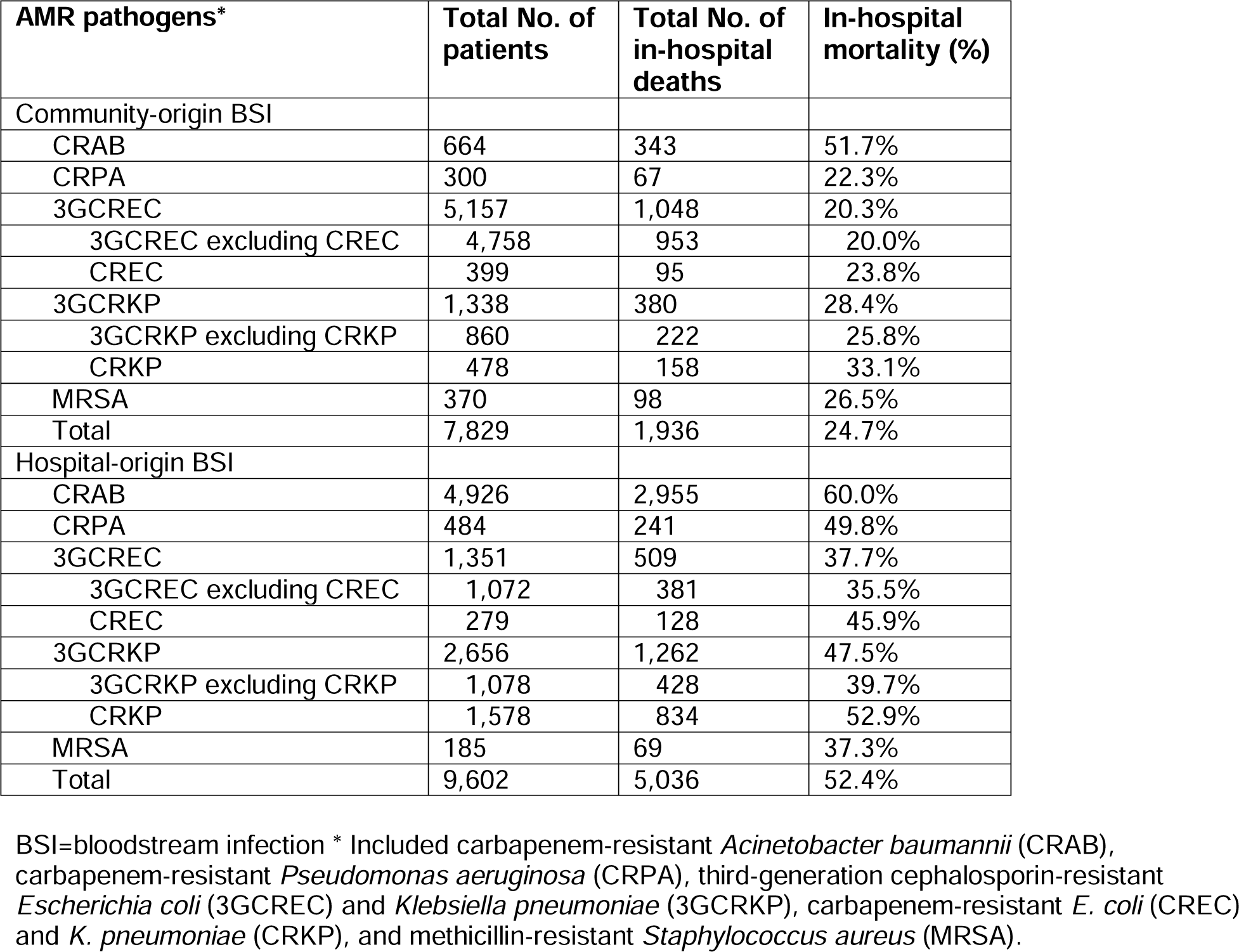
Total No. of cases and deaths following AMR BSI in 111 public referral hospitals in Thailand, 2022.

There were 9,567 patients with hospital-origin AMR BSI caused by the pathogens under evaluation. The most common cause of hospital-AMR BSI was CRAB, accounting 51.3% (4,926/9,602) of those patients. The total number of deaths following hospital-origin CRAB BSI was 2,955, representing 58.7% (2,955/5,036) of all deaths following hospital-origin AMR BSI caused by pathogens under evaluation.

### Factors associated with the frequency of community-origin 3GCREC BSI

The median observed frequency of community-origin 3GCREC BSI per 100,000 patients tested for community-origin BSI was 961 in Level A hospitals, 1,008 in Level S hospitals and 911 in Level M1 hospitals. In the multivariable regression model (Table 3), hospital level and bed size were not associated with the frequency of community-origin 3GCREC BSI (p=0.50 and 0.14, respectively). Hospital region was independently associated with the frequency (p<0.001). Hospitals in health regions 4 and 12 were associated the highest and lowest frequency, respectively, and the adjusted incidence rate ratio (aIRR) was about double (aIRR, 2.07; 95% confidence interval [CI] 1.53-2.80; Figure 1B). Rate of patients having BC taken per 1,000 patient-days was not included in the final model because of collinearity with the other variable for BC utilization. Percentage of patients having BC taken within the first two calendar days of hospital admission was negatively associated with the frequency of community-origin 3GCREC BSI (aIRR 0.88, 95%CI 0.82-0.94, per 5 percentage point change, p<0.001).

**Table 3.** Factors associated with the frequency of community-origin 3GCREC BSI per 100,000 patients tested for community-origin BSI.

| Factors | Crude incidence rate ratio (95% CI) | P value | Adjusted incidence rate ratio (95% CI) | P value |
| --- | --- | --- | --- | --- |
| A | 1.0 | 0.26 | 1.0 | 0.50 |
| S | 0.98 (0.82-1.17) |  | 1.07 (0.84-1.36) |  |
| M1 | 0.84 (0.68-1.05) |  | 0.98 (0.71-1.33) |  |
| Health regions |  |  |  |  |
| 1 | 1.33 (1.14-1.55) | <0.001 | 1.38 (1.03-1.85) | <0.001 |
| 2 | 1.77 (1.52-2.07) |  | 1.90 (1.40-2.56) |  |
| 3 | 1.81 (1.53-2.15) |  | 1.90 (1.35-2.67) |  |
| 4 | 1.94 (1.67-2.25) |  | 2.07 (1.53-2.80) |  |
| 5 | 2.04 (1.78-2.35) |  | 2.01 (1.52-2.65) |  |
| 6 | 1.94 (1.68-2.23) |  | 1.92 (1.37-2.68) |  |
| 7 | 2.28 (1.97-2.62) |  | 2.03 (1.47-2.80) |  |
| 8 | 1.71 (1.48-1.97) |  | 1.67 (1.26-2.22) |  |
| 9 | 1.20 (1.04-1.40) |  | 1.27 (0.92-1.75) |  |
| 10 | 1.23 (1.04-1.44) |  | 1.24 (0.89-1.73) |  |
| 11 | 1.40 (1.20-1.62) |  | 1.46 (1.10-1.94) |  |
| 12 | 1.0 |  | 1.0 |  |
| Hospital bed size (per 100 bed change)* | 1.02 (0.99-1.05) | 0.24 | 1.04 (0.99-1.09) | 0.14 |
| Gross provincial product* | 1.08 (1.01-1.16) | 0.034 | 0.99 (0.91-1.08) | 0.83 |
| Rate of patients having BC taken per 1,000 patient-days* | 0.83 (0.77-0.90) | <0.001 | - | - |
| Percentage of patients having BC taken within the first two calendar days of hospital admission (per 5 percentage point change)* | 0.83 (0.77-0.90) | <0.001 | 0.88 (0.82-0.94) | <0.001 |

\* The rate ratios for the four continuous variables denote the relative change in the frequency of community-origin 3GCREC BSI per 100,000 patients with the first BC taken within the first 2 calendar days of hospital admissions (i.e. patients tested for community-origin BSI). All four continuous variables were standardized by centering at the mean values and divided by the standard deviation, except that hospital bed size was divided by 100 and the percentage of patients tested for community-origin BSI by 5. The multivariable model consists of 5,157 patients with community-origin 3GCREC BSI from 522,919 patients tested for community-origin BSI from 111 hospitals.

There was between-hospital variation in the frequency of community-origin 3GCREC BSI in the final multivariable regression model (the estimated variance of the distribution of the random effects=0.066, p<0.001). The MRR was 1.28, representing the magnitude of between-hospital variation in the frequency of community-origin 3GCREC BSI. The median VPC was 0.71 (Figure S1 and S2), suggesting that, on average, 71% of the variation in the number of patients with community-origin 3GCREC BSI was due to differences between hospitals, while the remaining 29% was based on factors included in the final multivariable models.

Similar findings were also observed for the frequency of community-origin 3GCREC BSI per 100,000 admissions (Table S2), except that percentage of patients having BC taken within the first two calendar days of hospital admission was positively associated with the frequency of community-origin 3GCREC BSI per 100,000 admissions (aIRR 1.20, 95%CI 1.12-0.30, per 5 percentage point change, p<0.001).

### Factors associated with the frequency of hospital-origin CRAB BSI

The median observed frequency of hospital-origin CRAB BSI per 100,000 patients tested for hospital-origin BSI was 4,768 in Level A hospitals, 3,185 in Level S hospitals and 1,537 in Level M1 hospitals. In the multivariable regression model (Table 4), hospital level and bed size were independently associated with the frequency of hospital-origin CRAB BSI (p=0.010 and 0.001, respectively). Hospital region was not strongly associated with the frequency of hospital-origin CRAB BSI (p=0.096). The percentage of patients having BC taken after the first two calendar days of hospital admission was negatively associated with the frequency of hospital-origin CRAB BSI (aIRR 0.91, 95%CI 0.82-1.00, per 1 percentage point change, p=0.060).

**Table 4.** Factors associated with the frequency of hospital-origin CRAB BSI per 100,000 patients tested for hospital-origin BSI.

| Factors | Crude incidence rate ratio (95% CI) | P value | Adjusted incidence rate ratio (95% CI) | P value |
| --- | --- | --- | --- | --- |
| Hospital levels |  |  |  |  |
| A | 1.0 | <0.001 | 1.0 | 0.010 |
| S | 0.63 (0.49-0.79) |  | 0.90 (0.62-1.32) |  |
| M1 | 0.30 (0.22-0.41) |  | 0.56 (0.34-0.93) |  |
| Health regions |  |  |  |  |
| 1 |  | 0.40 | 1.33 (0.79-2.23) | 0.096 |
| 2 | 1.58 (0.86-2.92) |  | 2.11 (1.23-3.60) |  |
| 3 | 1.99 (1.00-3.97) |  | 2.40 (1.31-4.38) |  |
| 4 | 1.49 (0.83-2.67) |  | 1.99 (1.16-3.42) |  |
| 5 | 1.38 (0.81-2.36) |  | 1.69 (1.03-2.78) |  |
| 6 | 1.83 (1.05-3.17) |  | 1.93 (1.07-3.51) |  |
| 7 | 2.15 (1.10-4.17) |  | 2.38 (1.35-4.20) |  |
| 8 | 1.26 (0.71-2.25) |  | 1.57 (0.95-2.60) |  |
| 9 | 1.06 (0.56-2.03) |  |  |  |
| 10 | 1.11 (0.57-2.15) |  | 1.18 (0.67-2.08) |  |
| 11 | 1.49 (0.85-2.61) |  | 1.78 (1.07-2.98) |  |
| 12 | 1.54 (0.86-2.79) |  | 1.76 (1.05-2.96) |  |
| Hospital bed size (per 100 bed change)* | 1.16 (1.11-1.21) | <0.001 | 1.15 (1.06-1.24) | 0.001 |
| Gross provincial product* | 1.10 (0.98-1.23) | 0.12 | 1.03 (0.90-1.17) | 0.62 |
| Rate of patients having BC taken per 1,000 patient-days* | 0.99 (0.86-1.12) | 0.83 |  | - |
| Percentage of patients having BC taken after the first two calendar days of hospital admission (per 1 percentage point change)* | 1.16 (1.05-1.27) | 0.002 | 0.91 (0.82-1.00) | 0.060 |

\* The rate ratios for the four continuous variables denote the relative change in the frequency of hospital-origin CRAB BSI per 100,000 patients with the first BC taken after the first 2 calendar days of hospital admissions (i.e. patients tested for hospital-origin BSI). All four continuous variables were standardized by centering at the mean values and divided by the standard deviation, except that hospital bed size was divided by 100 and the percentage of patients tested for hospital-origin BSI by 1. The multivariable model consists of 4,926 patients with hospital-origin CRAB BSI from 115,516 patients tested for hospital-origin BSI from 111 hospitals.

There was between-hospital variation in the frequency of hospital-origin CRAB BSI in the final multivariable regression model (the estimated variance of the distribution of the random effects=0.184, p<0.001). The MRR was 1.51, representing the magnitude of between-hospital variation in the frequency of hospital-origin CRAB. The median VPC was 0.84 (Figure S3), suggesting that on average 84% of the variation in the number of patients with hospital-origin CRAB BSI was due to differences between hospitals. Figure S4 shows the differences in the frequency of hospital-origin CRAB BSI of each hospital within the same hospital level.

Similar findings were also observed for frequency of hospital-origin CRAB BSI per 100,000 admissions and per 100,000 patient-days at risk, except that percentage of patients having BC taken after the first two calendar days of hospital admission was positively associated with the frequency of hospital-origin CRAB BSI per 100,000 admissions (aIRR 1.15, 95%CI: 1.04-1.28, p=0.009, Table S3) and per 100,000 patient-days at risk of hospital-origin BSI (aIRR 1.09, 95%CI 0.99-1.20, p=0.092, respectively, Table S4).

## Discussion

This study demonstrates the regional differences in frequency of community-origin 3GCREC BSI in Thailand in 2022 after adjustment for the difference in hospital level, hospital bed size and rate of patients having BC taken. Our study highlights the high variation of the frequency of hospital-origin CRAB BSI between hospitals, suggesting that unadjusted factors at the hospital level (e.g. differences in case-mix, and infection prevention control (IPC) and antimicrobial stewardship (AMS) activities) are important.

Some health regions (e.g. 4, 5 and 7; Figure 1B) had relatively higher frequency of community-origin 3GCREC BSI than the others. Lack of association between hospital level and the frequency of community-origin 3GCREC BSI is consistent with the previous finding [8]. Our study did not observe an association between the local economy (as measured by GPP) and the frequency of community-origin 3GCREC BSI. Previous studies reported that regions, sex, age and underlying diseases are associated with the incidence rate of AMR *E. coli* BSI [18, 19]. Further studies are needed to evaluate whether these factors (using patient-level data) and other factors (e.g. antibiotic consumption at the provincial-level or regional-level) are associated with the frequency of community-origin 3GCREC BSI. Those insights would be crucial for formulating targeted interventions against community-acquired AMR infections in the country.

Association between hospital level and bed size and the frequency of hospital-origin CRAB BSI is consistent with the previous findings [8, 20]. This is likely because large referral hospitals tend to have a higher proportion of patients with severe conditions, or requiring complex surgery, prolonged intubation or urinary catheters, and a higher proportion of intensive care beds. Further studies are needed to evaluate and adjust for these factors using patient-level data. The substantial between-hospital variation for hospital-origin CRAB BSI may also be due to the differences in effectiveness of IPC and AMS activities implemented in each hospital. Due to the inability to adjust for differences in case-mix, we cannot conclusively determine whether hospitals with the highest frequency of hospital-origin CRAB BSI had less-effective IPC or AMS activities. However, the MoPH is promptly prioritizing these hospitals for immediate actions, including re-evaluation and reinforcement of IPC and AMS activities.

In line with previous studies [21, 22], our study shows that the under-utilization of BC could lead to higher observed frequency of AMR BSI per 100,000 tested patients and the lower observed frequency of AMR BSI per 100,000 admissions and per 100,000 patient-days. Possibly, hospitals which under-utilize BC tend to perform BC sampling primarily among patients with very severe infections or failure of empirical treatment. This would lead to a selection bias for culture positivity for AMR pathogens (resulting in a higher frequency of AMR BSI per 100,000 tested patients being observed) and to misdiagnoses of AMR BSI in patients without BC taken (resulting in a lower frequency per 100,000 admissions and per 100,000 patient-days being observed) [21].

Our study has multiple strengths. First, the number of hospitals included in the analysis was large. We also included a wide range of hospital levels and size. Second, we calculated and adjusted for BC utilization in the multivariable models. Third, we systematically estimated the magnitude of between-cluster variation [17, 23].

However, our study also has several limitations. First, we utilized summary data. Individual-level data such as sex, age and underlying diseases were not available for analysis. Using individual-level data and adjusting for patient-specific factors could reduce the VPC and MRR (i.e. between-cluster variation) [23]. Second, the origin of infection was only a proxy for community-acquired and hospital-acquired infection [24], and data of inter-hospital patient transfers were not available in our study. A proportion of patients with community-origin BSI, therefore, could be caused by nosocomial infections acquired at transferring hospitals. Third, mortality reported in this study was all-cause in-hospital mortality, and could be much lower than the true all-cause mortality because a preference to die at home is high in some regions in Thailand [25]. Fourth, the difference in the percentage of patients having BC taken could be due to the differences in case-mix. Further studies could additionally utilize antimicrobial prescription data to determine proportion of patients having a BC taken within ±1 calendar day of the day when a parenteral antibiotic was started at the study hospital and continued for at least four consecutive days [21, 22]. This could improve our understanding of differences in diagnostic use practice between hospitals and reinforce diagnostic stewardship activities in hospitals that underutilize BC among sepsis patients.

### Conclusions

Our findings highlight health regions where actions against community-acquired AMR infection, including reduction of antibiotic overuse in the community, should be considered. Our findings also highlight hospitals where actions against hospital-acquired AMR infection, including antimicrobial stewardship, infection control and diagnostic stewardship, should be prioritized.

## Acknowledgements

We gratefully acknowledge the laboratory team, IT team, antimicrobial stewardship team and infection control team of all public referral hospitals in Thailand for their participation in the activities of the Ministries of Public Health (MoPH), including utilization of the AMASS and submission of summary data to the MoPH.

## Transparency declarations

We declare that no competing interests exist.

## References

1. WHO. Global Action Plan on Antimicrobial Resistance2015. Available from: https://www.who.int/publications/i/item/9789241509763.

2. WHO. WHO report on surveillance of antibiotic consumption. 2016-2018 early implementation. 2019. Available from: https://www.who.int/publications/i/item/who-report-on-surveillance-of-antibiotic-consumption.

3. WHO. Global antimicrobial resistance and use surveillance systems | Country Profiles [Available from: https://www.who.int/data/gho/data/themes/topics/global-antimicrobial-resistance-surveillance-system-glass/glass-country-profiles.

4. Johnson AP, Muller-Pebody B, Budd E, Ashiru-Oredope D, Ladenheim D, Hain D, et al. Improving feedback of surveillance data on antimicrobial consumption, resistance and stewardship in England: putting the data at your Fingertips. T. 2017;72(4):953–6.

5. Ashiru-Oredope D, Cunningham N, Casale E, Muller-Pebody B, Hope R, Brown CS, et al. Reporting England’s progress towards the ambitions in the UK action plan for antimicrobial resistance: the English surveillance programme for antimicrobial utilisation and resistance (ESPAUR). J Antimicrob Chemother. 2023;78(10):2387–91.

6. Walter J, Haller S, Blank HP, Eckmanns T, Abu Sin M, Hermes J. Incidence of invasive meticillin-resistant Staphylococcus aureus infections in Germany, 2010 to 2014. Euro Surveill. 2015;20(46).

7. Lim C, Miliya T, Chansamouth V, Aung MT, Karkey A, Teparrukkul P, et al. Automating the Generation of Antimicrobial Resistance Surveillance Reports: Proof-of-Concept Study Involving Seven Hospitals in Seven Countries. J Med Internet Res. 2020;22(10):e19762.

8. Lim C, Hantrakun V, Klaytong P, Rangsiwutisak C, Tangwangvivat R, Phiancharoen C, et al. Frequency and mortality rate following antimicrobial-resistant bloodstream infections in tertiary-care hospitals compared with secondary-care hospitals. PLoS One. 2024;19(5):e0303132.

9. Srisuphan V, Klaytong P, Rangsiwutisak C, Tuamsuwan K, Boonyarit P, Limmathurotsakul D. Local and timely antimicrobial resistance data for local and national actions: the early implementation of an automated tool for data analysis at local hospital level in Thailand. JAC Antimicrob Resist. 2023;5(4):dlad088.

10. Veerasarn K, Yuthagovit S, Chailorrat A. Prevalence of Brain Tumor in Thailand from 2005 to 2014: Data from the National Health Security Office. J Med Assoc Thai. 2016;99 Suppl 3:S62–73.

11. Ministry of Public Health, Thailand. List of Healthcare Facilities under Health Administration Division, Ministry of Public Health, Thailand 2023. 2023. Available from: http://dmsic.moph.go.th/index/detail/9188.

12. Ministry of Public Health, Thailand. AMR Surveillance and Monitoring MoPH Dashboard 2023. Available from: https://app.powerbi.com/view?r=eyJrIjoiZGY0ZTcxNjMtYmNlZi00ODAyLWEzYjctYzBhOGRlM2ZlN2YxIiwidCI6IjM1YjAwOTJjLTM1MDYtNDliYS1iMWFjLTU0NmQwOTI1NzEyZCIsImMiOjEwfQ%3D%3D.

13. WHO. GLASS manual for antimicrobial resistance surveillance in common bacteria causing human infection. 2023. Available from: https://iris.who.int/bitstream/handle/10665/372741/9789240076600-eng.pdf

14. Sinto R, Lie KC, Setiati S, Suwarto S, Nelwan EJ, Djumaryo DH, et al. Blood culture utilization and epidemiology of antimicrobial-resistant bloodstream infections before and during the COVID-19 pandemic in the Indonesian national referral hospital. Antimicrob Resist Infect Control. 2022;11(1):73.

15. Teerawattanasook N, Tauran PM, Teparrukkul P, Wuthiekanun V, Dance DAB, Arif M, et al. Capacity and Utilization of Blood Culture in Two Referral Hospitals in Indonesia and Thailand. Am J Trop Med Hyg. 2017;97(4):1257–61.

16. National Statistical Office of Thailand. Statistical Yearbook Thailand 2023, 2023. Available from: https://www.nso.go.th/public/e-book/Statistical-Yearbook/SYB-2023.

17. Austin PC, Stryhn H, Leckie G, Merlo J. Measures of clustering and heterogeneity in multilevel Poisson regression analyses of rates/count data. Stat Med. 2018;37(4):572–89.

18. MacKinnon MC, McEwen SA, Pearl DL, Lyytikainen O, Jacobsson G, Collignon P, et al. Increasing incidence and antimicrobial resistance in *Escherichia coli* bloodstream infections: a multinational population-based cohort study. Antimicrob Resist Infect Control. 2021;10(1):131.

19. Laupland KB, Gregson DB, Church DL, Ross T, Pitout JD. Incidence, risk factors and outcomes of *Escherichia coli* bloodstream infections in a large Canadian region. Clin Microbiol Infect. 2008;14(11):1041–7.

20. Anugulruengkitt S, Charoenpong L, Kulthanmanusorn A, Thienthong V, Usayaporn S, Kaewkhankhaeng W, et al. Point prevalence survey of antibiotic use among hospitalized patients across 41 hospitals in Thailand. JAC Antimicrob Resist. 2023;5(1):dlac140.

21. Lim C, Hantrakun V, Teerawattanasook N, Srisamang P, Teparrukkul P, Sumpradit N, et al. Impact of low blood culture usage on rates of antimicrobial resistance. J Infect. 2021;82(3):355–62.

22. Sinto R, Lie KC, Setiati S, Suwarto S, Nelwan EJ, Karyanti MR, et al. Diagnostic and antibiotic use practices among COVID-19 and non-COVID-19 patients in the Indonesian National Referral Hospital. PLoS One. 2024;19(3):e0297405.

23. Austin PC, Wagner P, Merlo J. The median hazard ratio: a useful measure of variance and general contextual effects in multilevel survival analysis. Stat Med. 2017;36(6):928–38.

24. WHO. Global antimicrobial resistance surveillance system (GLASS) report: Early implementation 2017-2018. 2019. Available from: https://apps.who.int/iris/bitstream/handle/10665/279656/9789241515061-eng.pdf?ua=1.

25. Hongsuwan M, Srisamang P, Kanoksil M, Luangasanatip N, Jatapai A, Day NP, et al. Increasing incidence of hospital-acquired and healthcare-associated bacteremia in northeast Thailand: a multicenter surveillance study. PLoS One. 2014;9(10):e109324.

