## Supplementary file for "Frequency of antimicrobial-resistant bloodstream infections in Thailand, 2022"

- **Table S1. Characteristics of 111 public referral hospitals in Thailand, 2022, by hospital class**

| **Characteristics** | **Hospitals (N=111)** | - **Class A hospitals**   **(N=35)** | - **Class S hospitals**   **(N=49)** | - **Class M1 hospitals**   **(N=27)** |
| --- | --- | --- | --- | --- |
| Total No. of bed | 54,564 | 27,922 | 19,910 | 6,732 |
| Total No. of admissions | 3,916,766 | 1,992,737 | 1,471,408 | 452,621 |
| Total patient-days | 23,675,466 | 12,411,984 | 8,660,296 | 2,603,185 |
| Total No. of patients (de-duplicated) | 3,042,979 | 1,525,176 | 1,157,163 | 360,640 |
| Total No. of patients having BC taken* | 618,187 | 330,753 | 221,090 | 66,344 |
| Hospital characteristics |  |  |  |  |
| Bed count, median (IQR, range) | 420  (300-650,  150-1,387) | 773  (655-881,  512-1,387) | 407  (324-502,  215-592) | 258  (220-300,  150-350) |
| No. of admissions, median (IQR, range) | 27,539  (20,195-47,645, 9,722-103,085) | 53,143  (45,017-69,308, 32,647-103,085) | 27,121  (21,819-37,948, 15,902-52,791) | 16,382  (14,904-19,219, 9722-23,212) |
| Patient-days, median (IQR, range) | 174,638  (115,129-283,232, 40,648-686,395) | 346,390  (274,115-411,833, 201,087-686,395) | 156,675  (123,964-234,585, 65,733-351,543) | 93,484  (75,611-118,997, 40,648-174,204) |
| No. of patients, median (IQR, range) | 22,292  (15,708-36,313, 7,268-77,376) | 42,672  (34,873-51,688, 26,042-77,376) | 21,703  (18,192-30,045, 13,194-42,142) | 12,851  (11,539-14,551, 7,268-18,869) |
| No. of patients having BC taken, median (IQR, range) * | 4,516  (2,768-7,222,  941-14,945) | 9,449  (6,504-12,079,  5,234-14,945) | 4,291  (3,171-5,841, 1,329-8,707) | 2,596  (1,817-3,099,  941-4,243) |
| Percentage of patients having BC taken, median (IQR, range) ** | 20.0%  (16.1-22.9%,  6.2-34.6%) | 22.2%  (18.6-24.6%,  10.9-30.8%) | 19.7%  (16.4-21.7%,  6.2-34.6%) | 16.6%  (14.6-22.5%,  8.9-31%) |
| No. of patients having BC taken within first two calendar days of hospital admission, median (IQR, range) *** | 3,880  (2,405-6,336,  579-12,231) | 8,099  (5,511-9,977,  2,729-12,231) | 3,714  (2,581-5,100,  579-7,602) | 2,300  (1,555-2,782,  815-3,919) |
| Percentage of patients having BC taken within first two calendar days of hospital admission, median, median (IQR, range) *** | 17.3%  (13.1-20.2%,  3.2-31.6%) | 18.8%  (15.6-21.0%,  5.3-26.5%) | 16.5%  (13.6-19.3%,  3.2-31.6%) | 15.0%  (12.5-20.3%,  7.7-28.9%) |
| No. of patients having the first BC taken after the first two calendar days of hospital admission, median (IQR, range) *** | 815  (429-1,377,  148-6,730) | 1,657  (1,448-2,222,  857-6,730) | 730  (522-948,  160-1,916) | 363  (229-446,  148-530) |
| Percentage of patients having the first BC taken after the first two calendar days of hospital admission, median (IQR, range) *** | 3.4%  (2.6-4.0%,  0.8-10.9%) | 4.2%  (3.6-5.1%,  2.6-10.9%) | 3.3%  (2.7-3.8%,  0.8-5.8%) | 2.6%  (2.2-2.9%,  1.3-3.9%) |
| Rate of patients having BC taken per 1,000 patient-days | 26.4  (21.2-32.5,  8.7-48.6) | 27.8  (21.5-32.5,  13.9-38.4) | 26.1  (21.3-30.1,  8.7-42.7) | 25.5  (17.1-35.6,  15.1-48.6) |
| Hospital class** |  |  |  |  |
| Class A | 35 (31.5%) | 35 (100%) | - | - |
| Class S | 49 (24.3%) | - | 49 (100%) | - |
| Class M1 | 27 (44.1%) | - | - | 27 (100%) |
| Health regions |  |  |  |  |
| 1 | 10 | 3 | 3 | 4 |
| 2 | 8 | 2 | 4 | 2 |
| 3 | 5 | 1 | 4 | - |
| 4 | 10 | 3 | 4 | 3 |
| 5 | 15 | 4 | 7 | 4 |
| 6 | 12 | 6 | 3 | 3 |
| 7 | 6 | 2 | 3 | 1 |
| 8 | 11 | 2 | 6 | 3 |
| 9 | 7 | 4 | - | 3 |
| 10 | 6 | 2 | 4 | - |
| 11 | 12 | 3 | 6 | 3 |
| 12 | 9 | 3 | 5 | 1 |

BC=blood culture. *The AMASS deduplicated the data by selecting only the first BC culture per patient per surveillance period and per infection origin. **Hospital class is defined by the MoPH Thailand. Class A is Advanced-level referral hospital, Class S is Standard-level referral hospital, and Class M1 is Mid-level referral hospital. ***Some patients had more than one admission, of which one had the first BC taken within the first two calendar days of hospital admission and the other one had the first BC taken after the first two calendar days of admission.

- **Table S2. Factors associated with the frequency of community-origin 3GCREC BSI per 100,000 admissions**

| **Factors** | **Adjusted incidence rate ratio (95% CI)** | **P value** |
| --- | --- | --- |
| Hospital classes |  |  |
| A | 1.0 | 0.46 |
| S | 1.13 (0.88-1.46) |  |
| M1 | 1.06 (0.76-1.47) |  |
| Health regions |  |  |
| 1 | 1.25 (0.92-1.70) | <0.001 |
| 2 | 1.77 (1.29-2.43) |  |
| 3 | 1.84 (1.29-2.64) |  |
| 4 | 1.97 (1.44-2.70) |  |
| 5 | 1.93 (1.44-2.58) |  |
| 6 | 1.85 (1.30-2.64) |  |
| 7 | 1.88 (1.34-2.64) |  |
| 8 | 1.59 (1.18-2.14) |  |
| 9 | 1.21 (0.86-1.69) |  |
| 10 | 1.06 (0.74-1.50) |  |
| 11 | 1.47 (1.09-1.98) |  |
| 12 | 1.0 |  |
| Hospital bed count (per 100 bed change)* | 1.04 (0.99-1.10) | 0.088 |
| Gross provincial product* | 1.00 (0.92-1.10) | 0.94 |
| Percentage of patients having BC taken within the first two calendar days of hospital admission (per 5 percentage point change)* | 1.20 (1.12-1.30) | <0.001 |

* The rate ratios for the three continuous variables denote the relative change in the frequency of community-origin 3GCREC BSI per 100,000 admissions. All three continuous variables were standardized by centering at the mean values and divided by the standard deviation, except that hospital bed count was divided by 100 and the percentage of patients tested for community-origin BSI by 5. The multivariable model consists of 5,157 patients with community-origin 3GCREC BSI from 3,916,766 admissions from 111 hospitals.

- **Table 3S. Factors associated with the frequency of hospital-origin CRAB BSI per 100,000 admissions**

| **Factors** | **Adjusted incidence rate ratio (95% CI)** | **P value** |
| --- | --- | --- |
| Hospital classes |  |  |
| A | 1.0 | 0.006 |
| S | 0.89 (0.59-1.34) |  |
| M1 | 0.53 (0.31-0.91) |  |
| Health regions |  |  |
| 1 | 1.31 (0.75-2.27) | 0.16 |
| 2 | 2.13 (1.20-3.78) |  |
| 3 | 2.43 (1.27-4.65) |  |
| 4 | 2.08 (1.16-3.71) |  |
| 5 | 1.70 (1.06-3.82) |  |
| 6 | 2.01 (0.99-3.70) |  |
| 7 | 2.02 (1.10-3.73) |  |
| 8 | 1.49 (0.87-2.56) |  |
| 9 | 1.0 |  |
| 10 | 1.12 (0.60-2.06) |  |
| 11 | 1.72 (0.99-2.98) |  |
| 12 | 1.82 (1.04-3.18) |  |
| Hospital bed count (per 100 bed change)* | 1.15 (1.05-1.25) | 0.002 |
| Gross provincial product* | 1.01 (0.88-1.17) | 0.85 |
| Percentage of patients having BC taken after the first two calendar days of hospital admission (per 1 percentage point change)* | 1.15 (1.04-1.28) | 0.009 |

* The rate ratios for the three continuous variables denote the relative change in the frequency of hospital-origin CRAB BSI per 100,000 admissions. All three continuous variables were standardized by centering at the mean values and divided by the standard deviation, except that hospital bed count was divided by 100 and the percentage of patients tested for hospital-origin BSI by 1. The multivariable model consists of 4,926 patients with hospital-origin CRAB BSI from 3,916,766 admissions from 111 hospitals.

- **Table 4S. Factors associated with the frequency of hospital-origin CRAB BSI per 100,000 patient-days at risk of hospital-origin BSI**

| **Factors** | **Adjusted incidence rate ratio (95% CI)** | **P value** |
| --- | --- | --- |
| Hospital classes |  |  |
| A | 1.0 | 0.004 |
| S | 0.80 (0.54-1.17) |  |
| M1 | 0.48 (0.29-0.80) |  |
| Health regions |  |  |
| 1 | 1.33 (0.79-2.26) | 0.077 |
| 2 | 2.08 (1.20-3.59) |  |
| 3 | 2.63 (1.42-4.86) |  |
| 4 | 1.62 (0.93-2.82) |  |
| 5 | 1.53 (0.92-2.53) |  |
| 6 | 1.98 (1.08-3.64) |  |
| 7 | 2.48 (1.39-4.43) |  |
| 8 | 1.78 (1.06-2.99) |  |
| 9 | 1.0 |  |
| 10 | 1.30 (0.73-2.32) |  |
| 11 | 1.54 (0.91-2.60) |  |
| 12 | 1.88 (1.11-3.20) |  |
| Hospital bed count (per 100 bed change)* | 1.12 (1.03-1.21) | 0.008 |
| Gross provincial product* | 0.98 (0.53-1.12) | 0.73 |
| Percentage of patients having BC taken after the first two calendar days of hospital admission (per 1 percentage point change)* | 1.09 (0.99-1.20) | 0.092 |

* The rate ratios for the three continuous variables denote the relative change in the frequency of hospital-origin CRAB BSI per 100,000 patient-days at risk of hospital-origin BSI. Patients were considered at risk for hospital-origin BSI after they stayed in the hospital for more than two calendar days. All three continuous variables were standardized by centering at the mean values and divided by the standard deviation, except that hospital bed count was divided by 100 and the percentage of patients tested for hospital-origin BSI by 1. The multivariable model consists of 4,926 patients with hospital-origin CRAB BSI from 16,116,552 patient-days at risk of hospital-origin BSI from 111 hospitals.

- **Figure S1. Variance partition coefficient (VPC) for the final multilevel multivariable Poisson model evaluating factors associated with frequency of community-origin 3GCREC BSI per 100,000 patients tested for community-origin BSI**

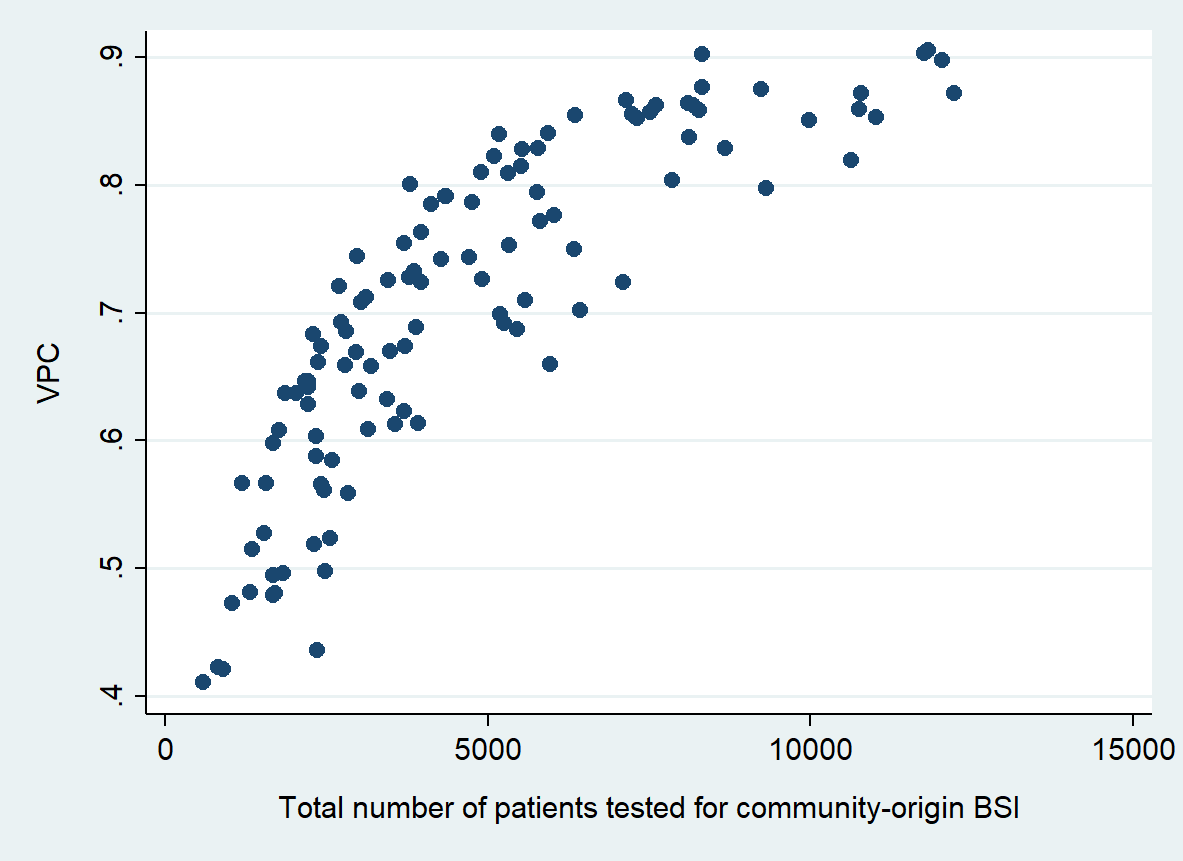

Footnote of Figure S1. The variation in the VPC was high and depended on the exposure (total number of patients tested for community-origin BSI). The median of the VPC was 0.71 (IQR 0.61-0.82, range 0.41-0.91)

- **Figure S2. Observed frequency of community-origin 3GCREC BSI among class A (S2A), S (S2B) and M1 (S2C) hospitals**

**S2A**

-
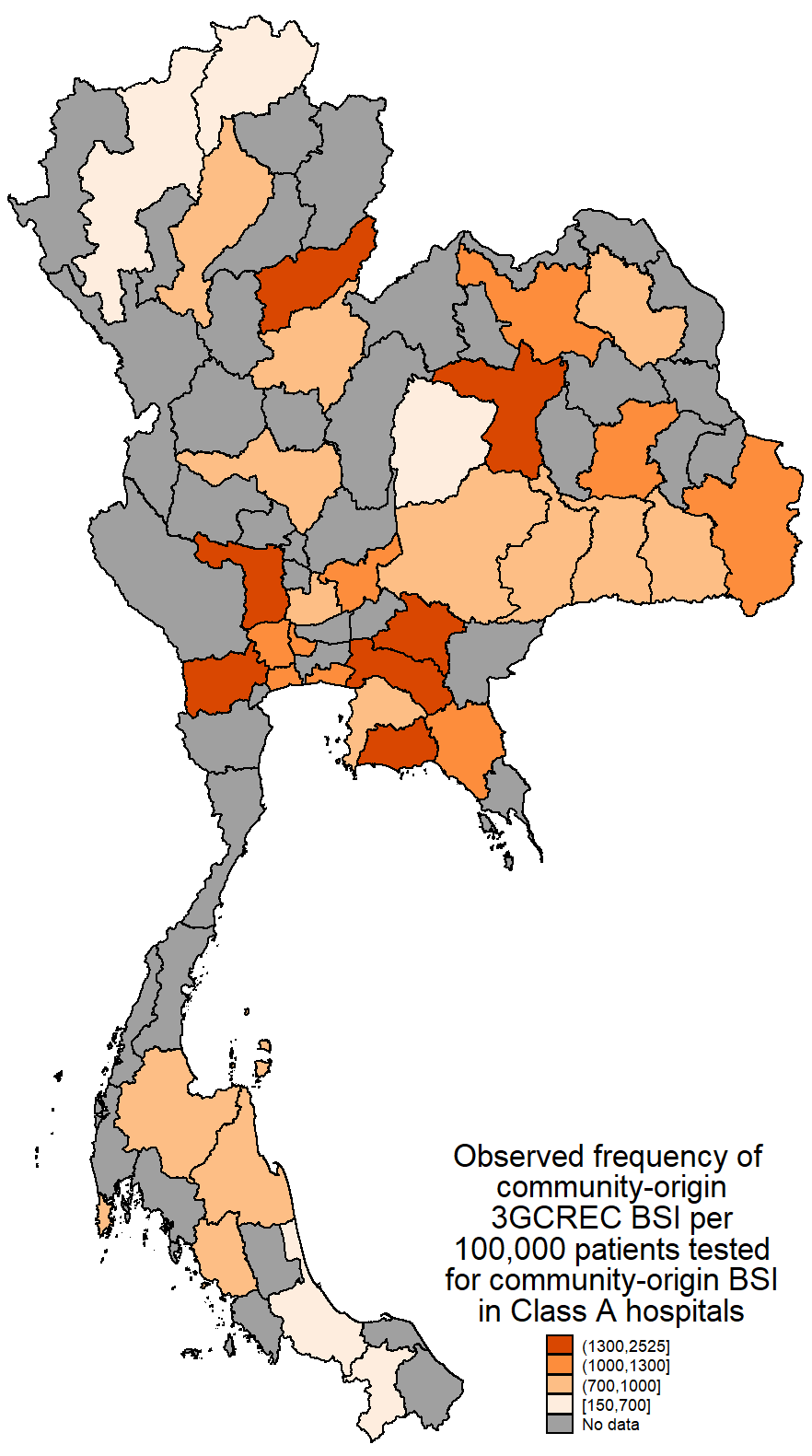

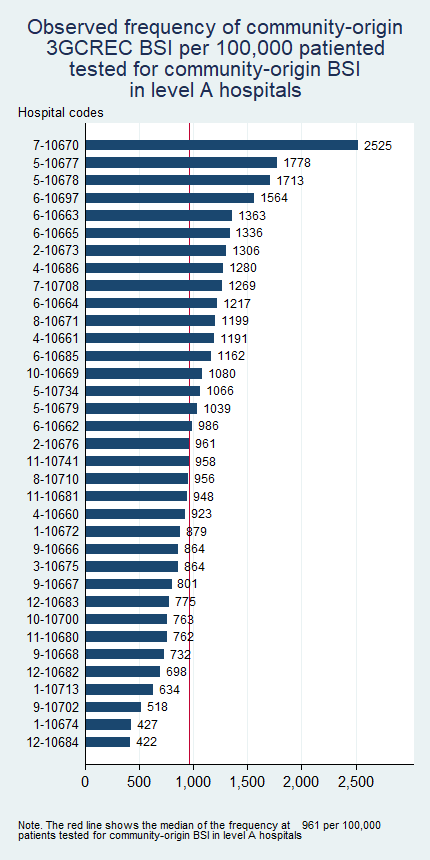

**S2B**

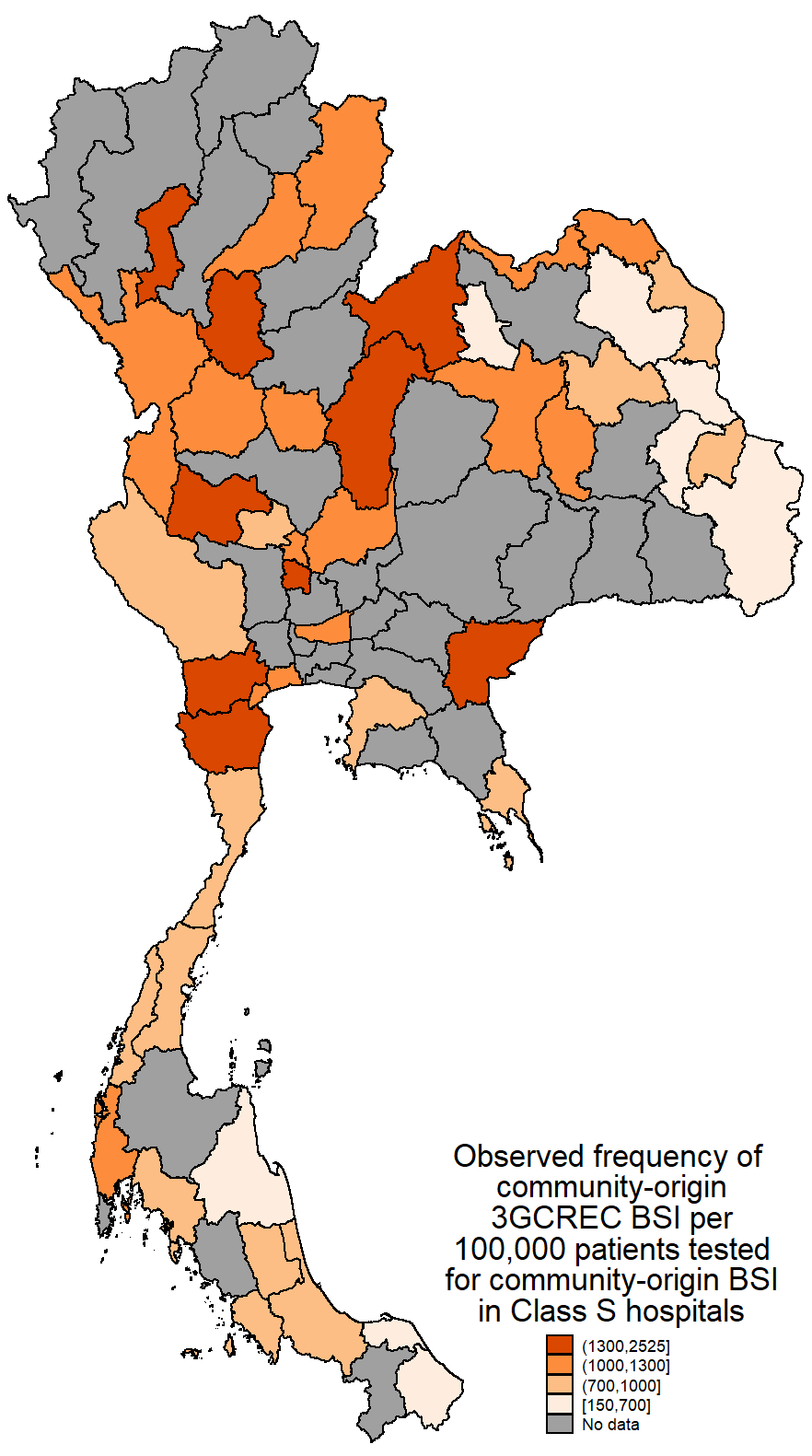

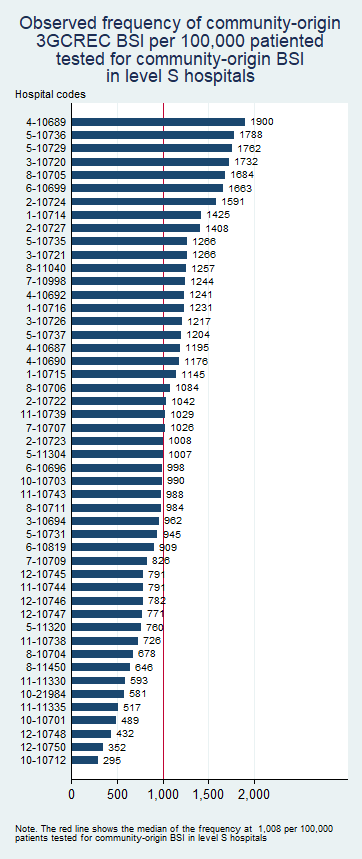

**S2C**

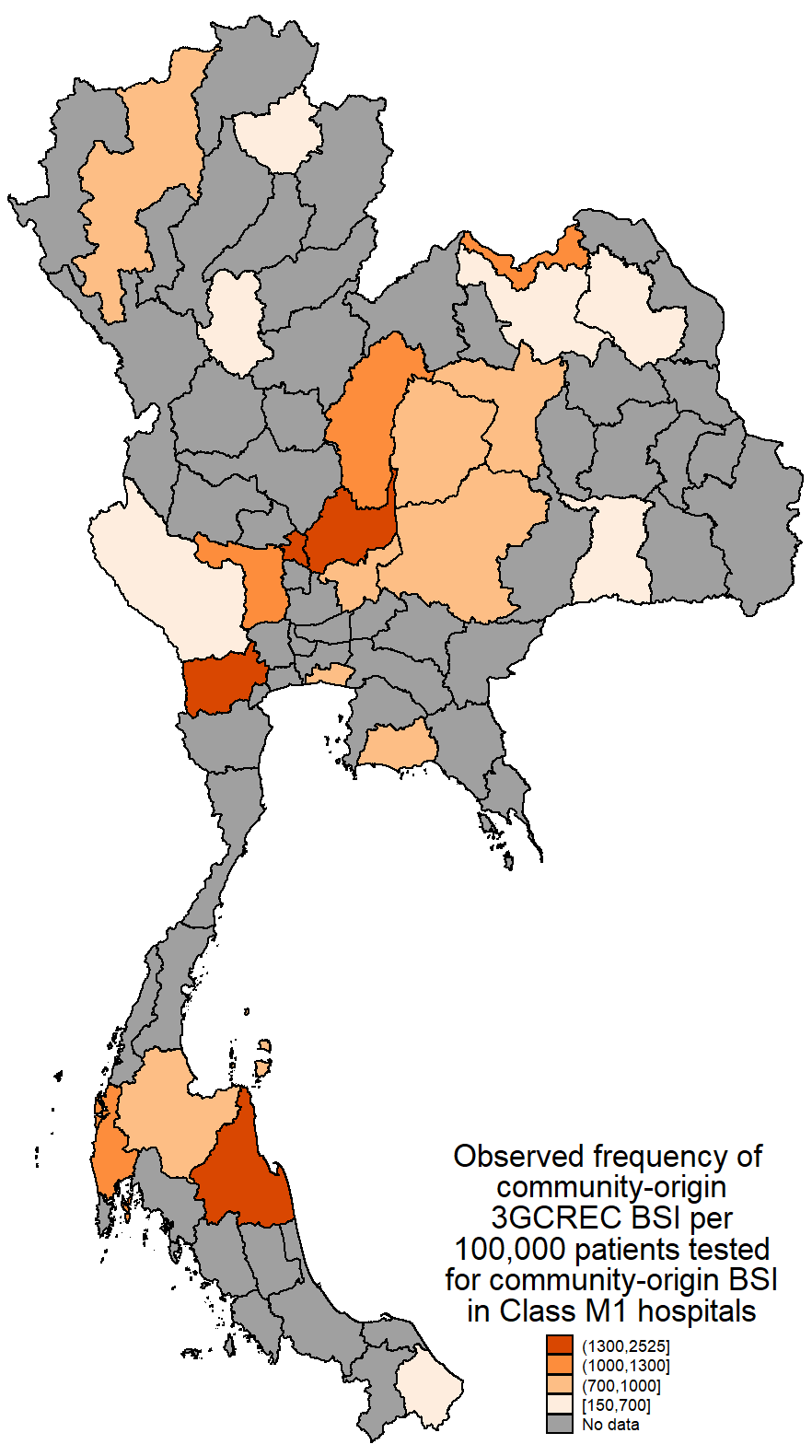

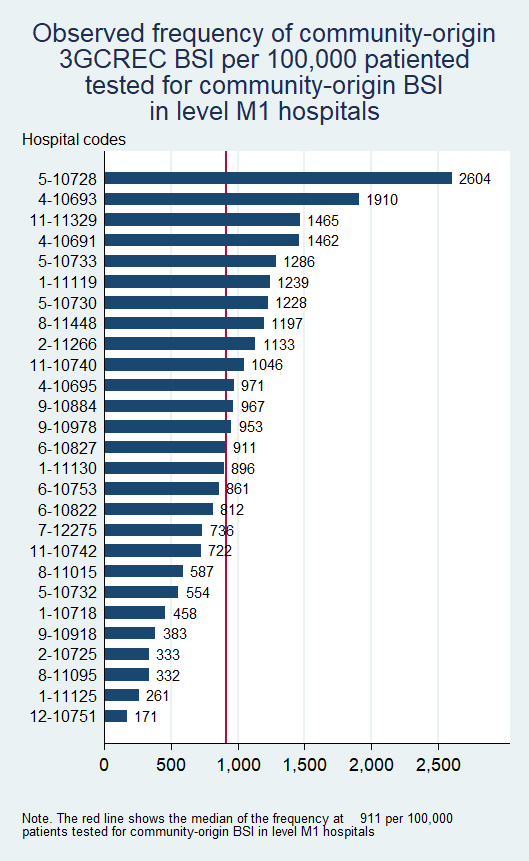

Footnote of Figure S2. In the provincial map, for provinces that had more than one hospital with similar hospital class, the total frequency was estimated based on the total number of observed cases per the total number of patients tested for community-origin BSI in the provinces.

- **Figure S3. Variance partition coefficient (VPC) for the final multilevel multivariable Poisson model evaluating factors associated with frequency of hospital-origin CRAB BSI per 100,000 patients tested for community-origin BSI**

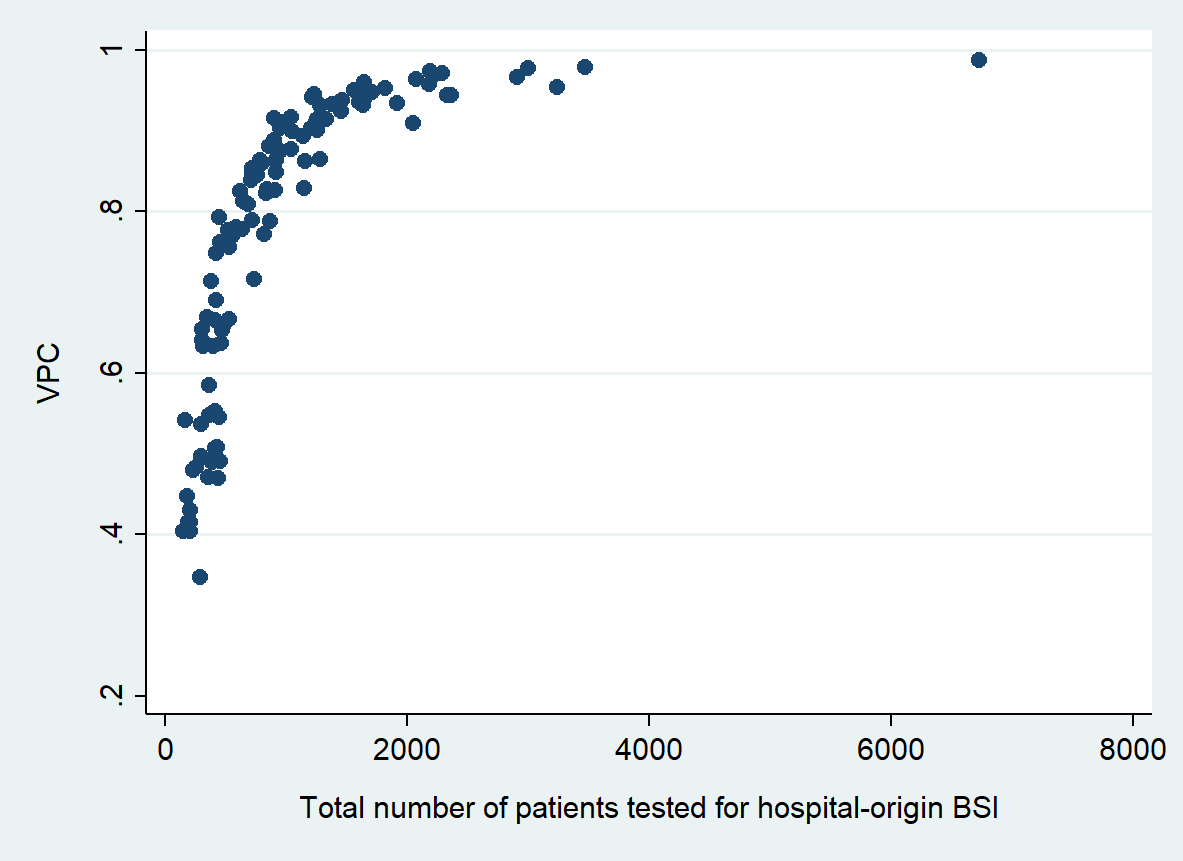

Footnote of Figure S3. The variation in the VPC was high and depended on the exposure (total number of patients tested for hospital-origin BSI). The median of the VPC was 0.84 (IQR 0.65-0.93, range 0.35-0.99).

- **Figure S4. Observed frequency of hospital-origin CRAB BSI among class A (S4A), S (S4B) and M1 (S4C) hospitals**

**S4A**

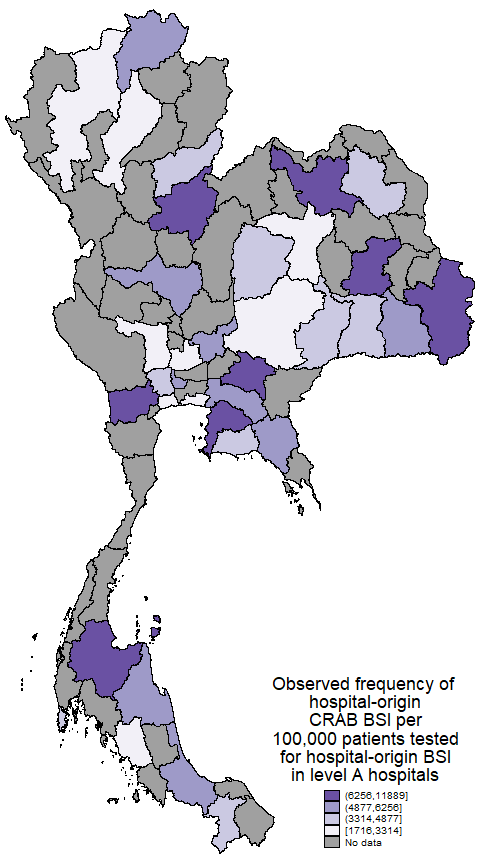

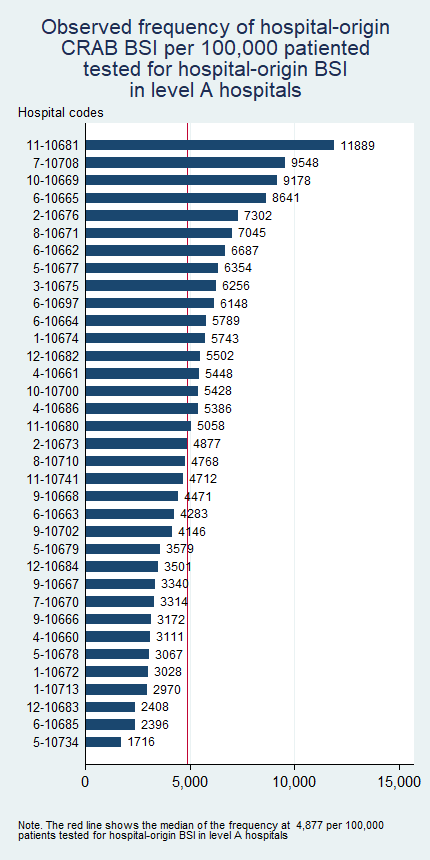

**S4B**

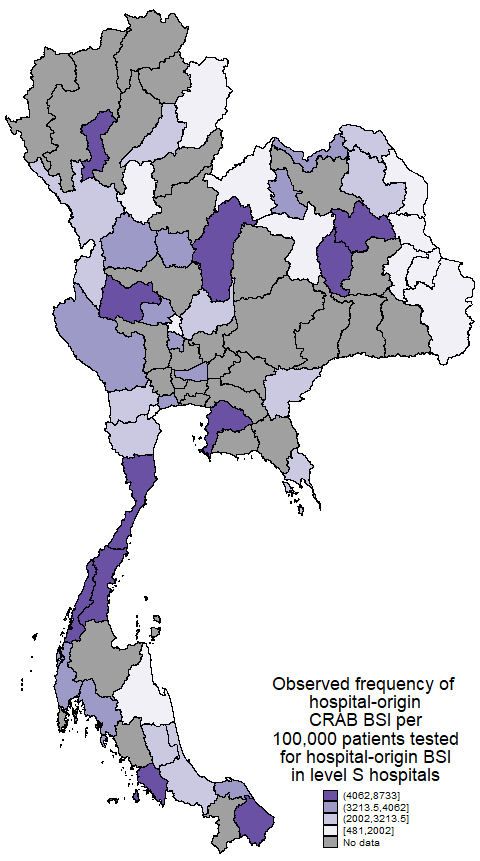

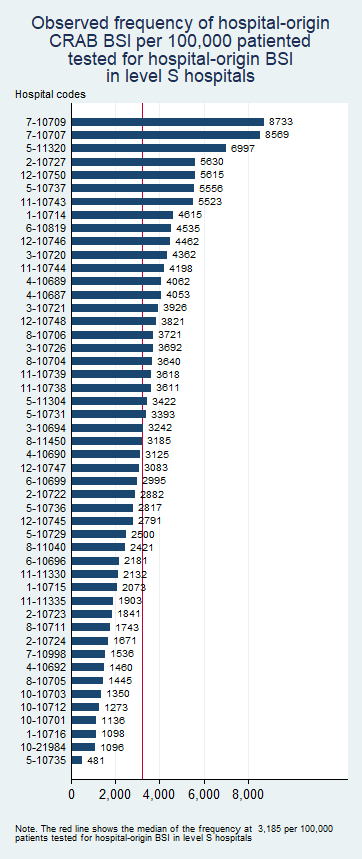

**S4C**

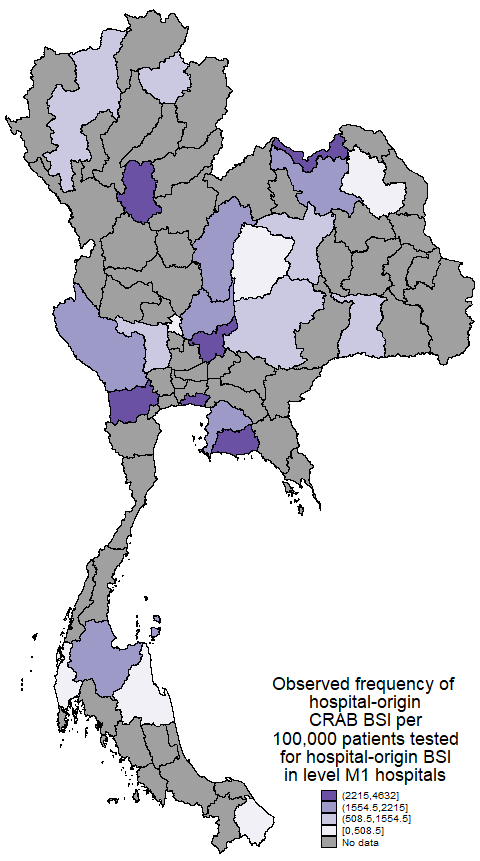

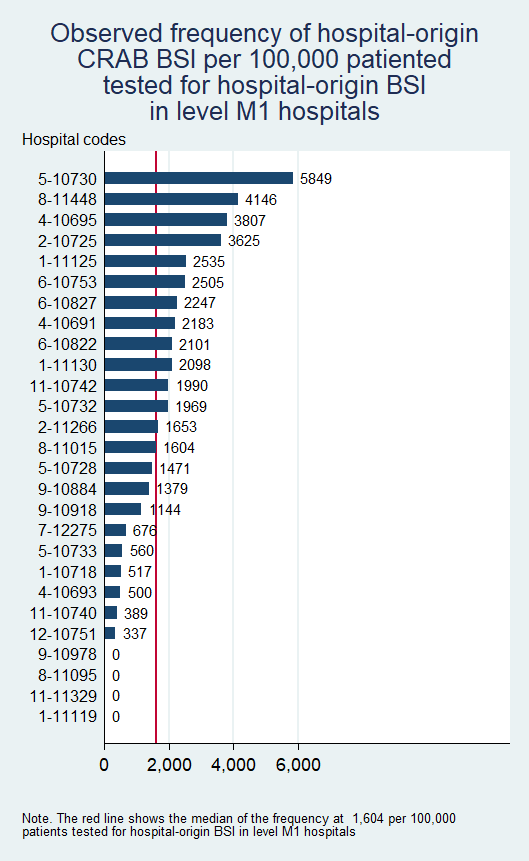

Footnote of Figure S4. In the provincial map, for provinces that had more than one hospital with similar hospital class, the total frequency was estimated based on the total number of observed cases per the total number of patients tested for community-origin BSI in the provinces.

**Appendix A**

**STATA code for fitting the multilevel multivariable Poisson regression model**

* Load the data

use TH_2022_111_hospitals, clear

******************************************************************************************

* Fit multilevel Poisson regression model with hospital-specific random effects

* for community-origin 3GCREC BSI per 100,000 patients tested for community-origin BSI

******************************************************************************************

mepoisson N_3GCREC_co i.Hospital_level ib12.Health_region Bed_count_S_100 GPP_2021_S Perc_tested_for_CAB_S_5 ///

, intmethod(mcaghermite) exposure(N_BCtested_for_CAB) ||Hospital_code:, irr

* Extract the variance of the random effects.

scalar tau2=_b[var(_cons[Hospital_code]):_cons]

scalar list tau2

* Extract the regression coefficients.

matrix beta_coefs = e(b)

matrix list beta_coefs

******************************************************************************************

* Compute the Median Rate Ratio

******************************************************************************************

scalar MRR = exp(sqrt(2*tau2)*invnorm(0.75))

scalar list MRR

******************************************************************************************

* ICC using exact calculations

******************************************************************************************

predict XB, xb

generate ICC=(exp(2*XB + 2*tau2) - exp(2*XB + tau2)) / (exp(2*XB + 2*tau2) - exp(2*XB + tau2) + exp(XB + tau2/2))

* Load the data

use TH_2022_111_hospitals, clear

******************************************************************************************

* Fit multilevel Poisson regression model with hospital-specific random effects

* for hospital-origin CRAB BSI per 100,000 patients tested for hospital-origin BSI

******************************************************************************************

mepoisson N_CRAB_ho i.Hospital_level ib9.Health_region Bed_count_S_100 GPP_2021_S Perc_tested_for_HAB_S_1 ///

, intmethod(mcaghermite) exposure(N_BCtested_for_HAB) ||Hospital_code:, irr

* Extract the variance of the random effects.

scalar tau2=_b[var(_cons[Hospital_code]):_cons]

scalar list tau2

* Extract the regression coefficients.

matrix beta_coefs = e(b)

matrix list beta_coefs

******************************************************************************************

* Compute the Median Rate Ratio

******************************************************************************************

scalar MRR = exp(sqrt(2*tau2)*invnorm(0.75))

scalar list MRR

******************************************************************************************

* ICC using exact calculations

******************************************************************************************

predict XB, xb

generate ICC=(exp(2*XB + 2*tau2) - exp(2*XB + tau2)) / (exp(2*XB + 2*tau2) - exp(2*XB + tau2) + exp(XB + tau2/2))
